# Strong effect of socioeconomic levels on the spread and treatment of the 2019 novel coronavirus (COVID-19) in China

**DOI:** 10.1101/2020.04.25.20079400

**Authors:** Zelong Zheng, Chloe Michelle, Xiangfeng Li

**Author notes:** Corresponding author: Chloe Michelle.

## Abstract

**Background:** Global response to the COVID-19 epidemic presents strengths and weaknesses in national and regional social governance capacities to address public health challenges. The emergence, detection, spread, treatment and containment of infectious diseases shows the considerable political and economic impacts in a highly interconnected world. We aimed to estimate the effects of socioeconomic levels on the spread and treatment of COVID-19 in China. **Methods** We obtained daily COVID-19 cases at a city level in China. We used migration data from the major cities in Hubei Province, and macroeconomic data at city and province levels. We obtained social management measures in response to COVID-19 outbreak. We assessed the association between measures, migration and COVID-19 spread, and the association between socioeconomic levels and COVID-19 treatment capacity.

**Findings:** On January 1, 2020, COVID-19 spread that affected by management measures and migration started across China. After Wuhan lockdown, the case number reached peak in 12 days, and COVID-19 outbreak was basically contained in China in four weeks due to intensive measures. Guangdong, Jiangsu and Zhejiang Provinces showed the most excellent COVID-19 treatment capacities. Socioeconomic levels in these provinces ranked top in China. Guangdong achieved the largest decline in severe case rate by 22.1%. Jiangsu had the lowest average rate of severe cases (1.7%) and zero death. Among the regions with top case number, Zhejiang showed the highest rate of cured cases on confirmed cases (96.3%), the lowest average rate of severe cases (7.7%), and one death. The COVID-19 treatment capacities were strongly affected by regional economics and measures on control, detection and treatment.

**Interpretation:** Socioeconomic levels had strong effect on the spread and treatment of COVID-19 in China. Further investigations are needed on the effectiveness of Chinese measures and the effects of socioeconomic levels on COVID-19 treatment outside China.

**Fund:** None

Research in context

**Evidence before this study:** We searched PubMed for articles published in any language up to April 24, 2020, with the search terms “COVID-19 AND (socioeconomic OR measure) AND (spread OR treatment)”. We identified 334 articles. Some researchers are dedicated to debating the effect of social management measures on the spread of COVID-19 epidemic. All previous studies focused on the effect of the individual measure on COVID-19 spread over time. We identified several mathematical modelling studies exploring the effect of management measures, mainly focusing on Wuhan lockdown in China, on COVID-19 spread. However, social management measures not only involve prevention and control of virus spread, but also virus detection and patient treatment. No study used methods that would allow the assessment of effect of several management measures on the spread, detection, and treatment of COVID-19 at various time milestones over the entire course of COVID-19 outbreak. Some scholars advocated that health equity cannot be ignored to contain the global COVID-19 epidemic. They did not provide epidemical and economic data analysis to assess the effect of socioeconomic gradients in health at individual or regional levels. No study estimated the effects of socioeconomic levels on national and regional COVID-19 treatment.

**Added value of this study:** We found that on January 1, 2020, COVID-19 spread that affected by management measures and migration started across China. After Wuhan lockdown, COVID-19 outbreak was basically contained in China in four weeks due to intensive measures. The intensive measures mainly include movement restriction, wearing masks in public, nationwide joint prevention and control at a community level, four early strategies, and information disclosure. We, for the first time, estimated the effect of socioeconomic levels on spread and treatment of COVID-19 in China. The management measures, including Fangcang shelter hospitals, medical assistance nationwide, and continuously updated diagnosis and treatment plan for COVID-19, greatly improved COVID-19 treatment capacities in China, particularly in Hubei Province. The COVID-19 treatment capacities were strongly affected by regional economics and measures on control, detection and treatment.

**Implications of all the available evidence:** The Chinese experience provides important insights into how to design effective management strategies of COVID-19 or other epidemic. Further efforts are needed on the effectiveness of Chinese management measures and the effects of socioeconomic levels on COVID-19 treatment outside China.

## 1. Introduction

Starting in December 2019, the 2019 novel coronavirus (COVID-19) broke out in Wuhan City, Hubei Province of China. From 31 December 2019 through 3 January 2020, a total of 44 cases with novel coronavirus pneumonia (NCP) of unknown etiology were reported to the World Health Organization (WHO) by the Chinese health authorities^1–3^. Confirmed cases were consecutively reported in 34 provinces, municipalities, and special administrative regions in China^4^. As of 20 April 2020, there have been 8,4201 confirmed cases of COVID-19 infection and 4642 deaths in China. Outside China, there have been 2,157,577 cases and 147,909 deaths in at least 215 countries and regions^5^. The COVID-19 pandemic is sweeping the world and delivers array of cybersecurity challenges, i.e., overload the healthcare system and disrupt the socioeconomic system^6–8^. The global response to the COVID-19 epidemic presents strengths and weaknesses in national and regional social governance capacities to address public health challenges. The emergence, detection, spread, treatment and containment of infectious diseases shows the considerable political and economic impacts in a highly interconnected world^9^. Understanding how the socioeconomic levels respond to the spread and treatment of COVID-19 is of great significance for the success of combating the coronavirus epidemic.

Social and economic circumstances affect the spread and treatment of infectious diseases. Conventionally, social governance measures, such as travel ban, social distancing and gathering reduction, play a major role in preventing and controlling the rapid spread of the epidemic in response to public health emergencies^10^. COVID-19 continues to immediately spread through migration-this was the case in Wuhan of China at the beginning of the pandemic and is now the cases all over China and the World^5,11^. Population migration and mobility is linked to increases in economic growth, wages, income and innovation as well as virus spread^12,13^. The lockdown of cities was adopted by many countries in succession after Wuhan was closed on 23 January 2020^14^. This measure aimed to mitigate the spread of COVID-19 and limit the number of patients that health systems have to manage. The most recent studies show that approximately 15–20 per cent of patients with COVID-19 require hospitalisation and six per cent require intensive care for a duration of between 3 and 6 weeks. Confirmation of the COVID-19 diagnosis requires laboratory and/or medical imaging capabilities that are only available in reference structures, like teaching hospitals^15^. Even in wealthy nations, U.S. and EU, they have strong, well-funded health services and the hospital system was quickly overwhelmed by the rapid increase in COVID-19 cases. Many poorer nations in Africa and Asia, such as Pakistan and Iraq, are facing larger difficulties^3^. These countries are equipped with under-developed infrastructure, inadequate pathogen detection capability and poor health care service, and they cannot afford the fewer patients in comparison with developed nations. The disruption of the health care systems led to a low rate of cure, high severity and mortality rates of the COVID-19 patients^7^. Furthermore, COVID-19 pandemic is likely to cause the disruption of basic medical services and emergency facilities, the de-prioritisation of treatment for other life-threatening diseases, conditions and for other chronic infectious diseases (e.g., cardiovascular and cerebrovascular diseases, tuberculosis and malignancy) everywhere but especially in some developing economies, where the health system is already fragile^16^. Thus, improved understanding of the spread and treatment capacities of COVID-19 that affected by socioeconomic levels is crucial to examining the effectiveness of control interventions.

In this study, we aim to estimate the effects of socioeconomic levels on the spread and treatment of the COVID-19 in China. The spatial and temporal patterns of the spread and treatment of the COVID-19 at city and province levels across China were identified. The effects of social management measures and migration on COVID-19 spread, and the effects of management measures and economic levels on COVID-19 treatment were assessed.

## 2. Materials and Methods

### 2.1 COVID-19 case data

From December 31, 2019, the centers of disease control (CDC)s at all levels in China jointly launched the COVID-19 investigation. COVID-19 was identified as a statutory B infectious disease in China on January 20, 2020. Legally, all cases were required to be reported immediately through the Infectious Disease Information System (IDIS). Individual case information was submitted into the system by local hospitals and CDC personnel who investigated and collected possible exposure information. All case records have personal identification numbers, and all cases are not duplicated in the system. We collected the COVID-19 cases on a daily basis that reported by the CDCs at all levels as of March 4, 2020, in the cities of 31 provinces, municipals and autonomous regions in Mainland China. These data and management measures were obtained from the websites of local health commissions. All cases were included in this study, and the sampling of a predetermined sample size was not required, and the case inclusion criteria was not considered ^17^.

Terms with respect to COVID-19 cases in China were defined based on treatment plan of COVID-19 that issued by Chinese authorities^17,18^. If the case has been in close contact with the Huanan Seafood Market, who was identified as an exposure linked to the Huanan Seafood Market. Symptom severity of the cases was classified as mild, severe, and critical. Mild cases were not our focus. We focused on confirmed cases, cured cases, and severe (i.e., severe and critical) cases. Suspected cases were determined based on the symptoms (e.g., fever, cough, fatigue and diarrhea) and exposures (if the patient lived or traveled in Wuhan, or had close contact with a person who has been to Wuhan recently, who was identified as Wuhan related exposure) for clinical diagnosis. Confirmed cases referred to suspected cases with positive results of nucleic acid testing through respiratory tract samples (e.g., throat swabs). Clinically diagnosed cases were those suspected cases with pneumonia imaging features (only applicable in Hubei Province). Cured cases referred to the cases that were cured and discharged. Severe cases referred to the cases that manifest dyspnea with respiratory frequency ≥30 / min, blood oxygen saturation ≤93%, PaO_2_ / FiO_2_ ratio <300, and / or lung infiltration> 50% within 24 to 48 hours. Critical cases referred to those cases with respiratory failure, septic shock and / or multiple organ dysfunction / failure.

### 2.2 Socioeconomic data

Data of migration from Wuhan and the other cities in Hubei Province on a daily basis were obtained from Qianxi Baidu website (https://qianxi.baidu.com/), ranging from January 1 to January 28 in 2019 and 2020. The top 20 cities in the migration size were assessed.

Macroeconomic data were collected from two levels: city and province level. At the province level, the data from the 31 provinces, municipalities and autonomous regions outside Hubei Province were obtained from China health statistics yearbook in 2019, and comprised the indicators of population, total health expenditure (billion Yuan), gross domestic product (GDP) (billion Yuan), total health expenditure in GDP (%), No. of hospitals, No. of top hospitals, No. of doctors, No. of nurses, No. of beds in general hospitals, and public expenditure (billion Yuan). At the city level, the data were obtained from China city statistics yearbook in 2019, and were available for the 292 cities outside Hubei Province. These data comprised the indicators of population, GDP (billion Yuan), No. of hospitals, No. of doctors, No. of hospital beds, and public expenditure (billion Yuan). The statistical indicators of macroeconomic and medical resource data at different levels were different. Spatial patterns of the indicators representing macroeconomic levels at city and province levels in China were illustrated using Arcgis Desktop 10.2 (Figure 7, 8 and 9). As the COVID-19 outbreak overloaded the health care system in Hubei Province, and a total number of 3,8478 health care workers in other provinces, municipalities and autonomous regions of China assisted Hubei (Chinese Health commission news). The treatment capacity in Hubei Province is difficult to be evaluated. Therefore, Hubei Province was excluded to estimate the effects of regional socioeconomics on the COVID-19 treatment. Spatial pattern of the number of health care workers in other provinces, municipalities and autonomous regions of China that assisted Hubei were illustrated using Arcgis Desktop 10.2 (Figure 6).

### 2.3 Social management measures in response to COVID-19 epidemiologic pattern

Social management measures in response to COVID-19 epidemiologic patterns were illustrated in Figure 1. On December 31, 2019, a cluster of pneumonia cases with unknown etiology were reported in Wuhan, Hubei province, China, which attracted great attention of health authorities. Wuhan Health Commission (WHC) declared novel coronavirus pneumonia (NCP) outbreak, and National Health Commission (NHC) China and Chinese CDC participated in investigation and response. The epidemiologic investigation pointed out that the case infection may be associated with exposures in Huanan Seafood Market in Wuhan. On January 1, 2020, Huanan Seafood Market was closed. On January 8, 2020, the pathogen causing the viral pneumonia among affected individuals was identified as a new coronavirus COVID-19. On January 15, 2020, China CDC started the public health emergency response level to Level 1 (the highest level). On January 20, 2020, COVID-19 was identified as a statutory B infectious disease in China. Human to human transmission was confirmed by Zhongnanshan, MD. Wuhan is located in central China and has a wide range of transportation links, including airplanes, trains, interstate buses, and private transportation. On January 23, 2020, Wuhan was closed to prevent and control COVID-19 spread. On January 24–28, 2020, the other cities in Hubei province were closed in succession. On January 27, 2020, Lunar New Year national holiday was extended. On January 29, 2020, 31 provinces, municipalities and regions in China started primary response to major public health, and return to work and school across China was delayed. On February 13, 2020, the number of clinically diagnosed cases of COVID-19 was included in Hubei Province, and the number of confirmed cases increased sharply. On February 17, 2020, workers started to return to work nationwide outside Hubei. Starting on February 24, 2020, the response levels of public health downgraded outside Hubei.

**Figure 1.**
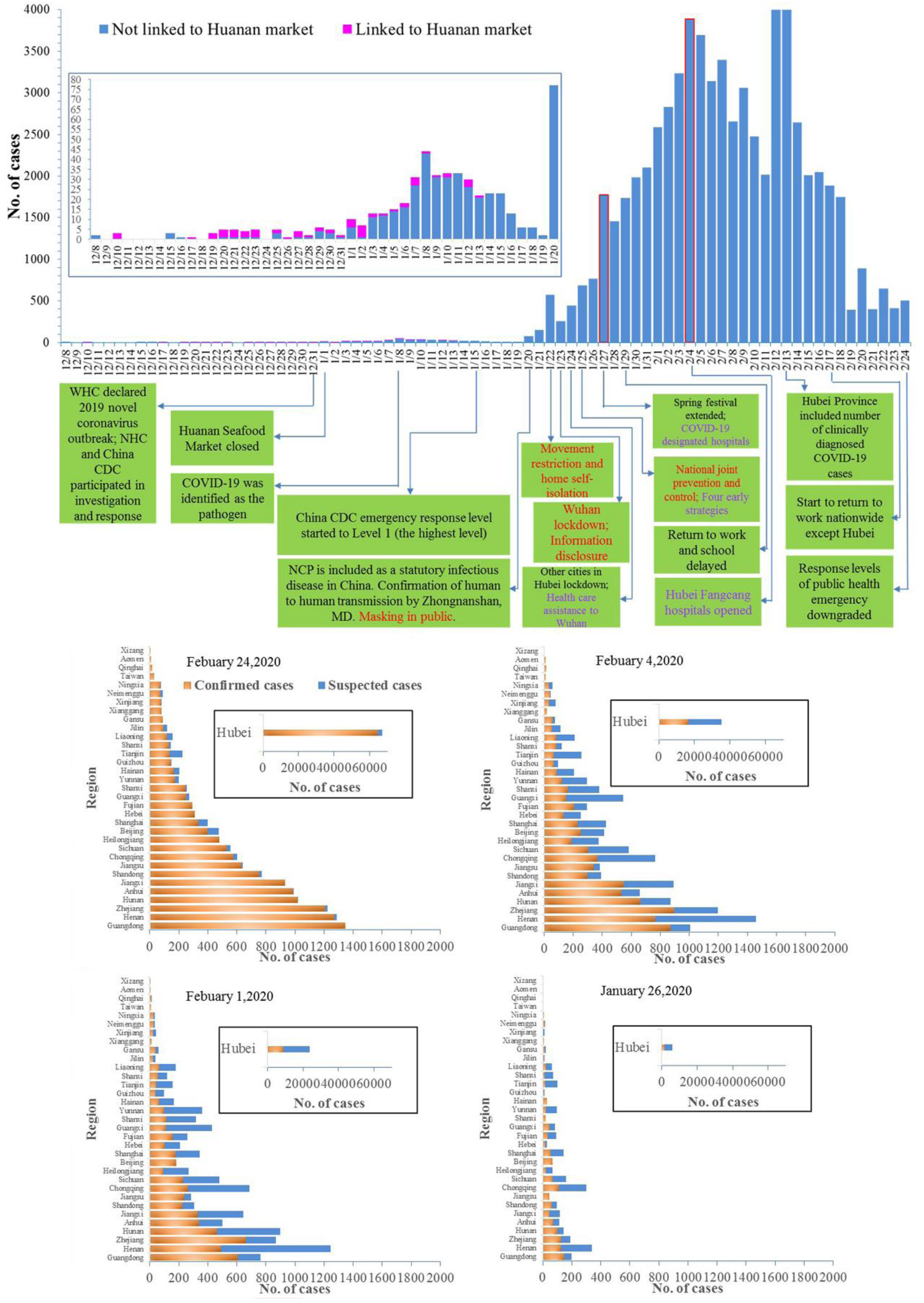
Epidemiologic patterns corresponding to milestones of confirmed and suspected cases of COVID-19 in China. The number of daily confirmed cases that were linked (blue) and unlinked (pink) to Huanan Seafood Market were plotted by the date of diagnosis. The time ranged from December 8, 2019, the date of case onset to February 24, 2020, the date of notable case decrease. The dates were used as a timeline of primary social management measures in response to COVID-19 outbreak in China. Social management measures taken by the Chinese authorities are shown in green boxes. The incidence of COVID-19 cases was few as of January 20, 2020, and these cases are indicated in the inset. The outlines of two peaks, January 27 and February 4, 2020 were highlighted in red. Social management measures on COVID-19 spread were highlighted in red, and the measures on COVID-19 treatment were highlighted in purple. At four key time nodes of COVID-19 cases, the number of confirmed (blue) and suspected (orange) cases in the 34 provinces (including Hubei), municipalities and autonomous regions were illustrated. (This figure was modified based on Wu and McGoogan ^17^)

Chinese government adopted intensive measures to contain COVID-19. i.e., Wuhan lockdown, movement restriction and home self-isolation, masking in public, national joint prevention and control, information disclosure, timely and sufficient COVID-19 detection. Masking in public is to protect oneself from infection. Since COVID-19 was proved to be transmitted mainly through droplets, wearing a mask is effective in preventing virus transmission. National joint prevention and control was launched down to the community level. Information transparency was guaranteed. i. e., real-time announcement of the number of confirmed and suspected cases every day in each city across China. COVID-19 was detected timely and quantitatively. Nucleic acid detection was carried out in all close contact including those without symptoms. Four early strategies were suggested: early protection (social distancing), early detection, early diagnosis, and early isolation. Management measures targeting COVID-19 treatment comprised: four early strategies, Fangcang shelter hospital construction, health care assistance from other regions of China to Hubei, and determination of designated hospitals for the COVID-19 treatment.

### 2.4. Statistical analysis

Epidemiologic patterns of COVID-19 outbreak in China were identified at the city and province levels on a daily basis. The outbreak period ranged from December 8, 2019, the date of case onset to February 24, 2020, the date of notable case decrease. The number of daily confirmed cases in China that were linked and unlinked to Huanan Seafood Market were plotted by the date of diagnosis. At four key time nodes of COVID-19 outbreak, January 27, February 1, February 4, and February 24, 2020, the number of confirmed and suspected cases in the 34 provinces (including Hubei), municipalities and regions were illustrated (Figure 1). Spatial patterns of confirmed cases at a city level across China on January 27, February 4 and March 4, 2020 were presented using Arcgis Desktop 10.2 (Figure 2).

**Figure 2.**
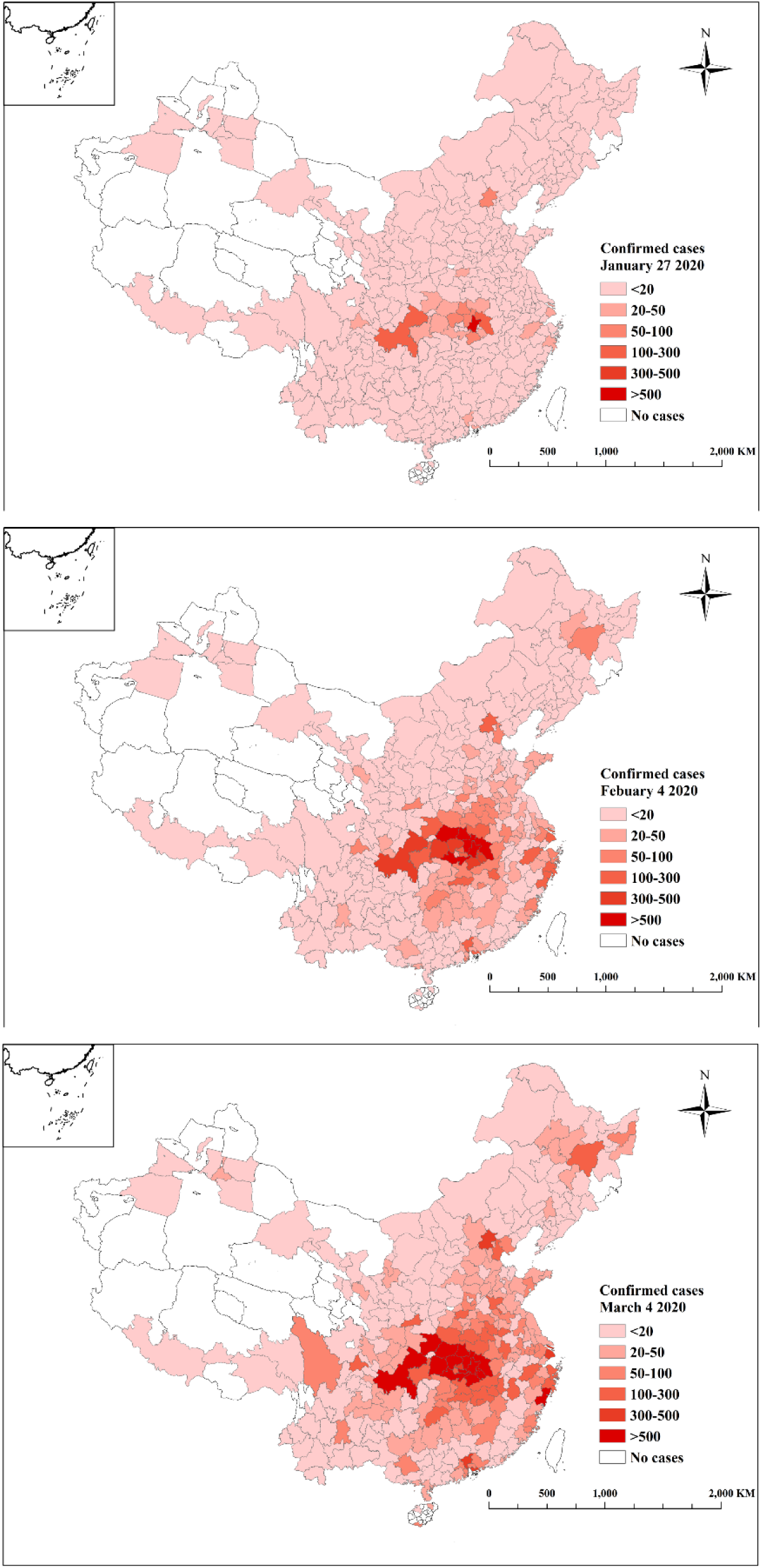
Spatial patterns of confirmed cases at a city level across China on January 27, February 4 and March 4, 2020

To assess the effects of management measures on COVID-19 spread, COVID-19 epidemiologic curve was made by the dates of the milestones that were associated with management measures. The impact of management measures on COVID-19 related migration in Wuhan, Huanggang, Jingzhou and Xiaogan cities in Hubei province were examined.

The relationship between migration from these cities and confirmed cases of COVID-19 in China (outside Hubei province) was examined. Daily migration scale index from these cities from January 1 to 28 in 2019 and 2020, respectively were calculated and compared. We selected this period because a significant increase in migration size occurred in Wuhan in 2020 in comparison with that in 2019. The management measures responded COVID-19 outbreak in Wuhan that may cause the abnormal increase in migration. Specifically, On January 1, 2020, The Huanan Seafood Market was closed because of COVID-19 outbreak. On January 8, 2020, the pathogen was identified as COVID-19. On January 15, 2020, China CDC started the public health emergency response level to Level 1. On January 20, 2020, COVID-19 was identified as a statutory B infectious disease in China, and human to human transmission was first confirmed. On January 23, 2020, Wuhan was closed. On January 24–28, 2020, other cities in Hubei province were closed in succession. These dates were the milestones when significant changes in the time series of migration scale index for Wuhan were identified. We thus divided the period, January 1 to 28, into five sub-periods: January 1 to 8, January 9 to 15, January 16 to 20, January 21 to 23 and January 24 to 28. The correlations between migration scale index in Wuhan, Huanggang, Jingzhou and Xiaogan cities in these five sub-periods and confirmed cases in China (outside Hubei province) on January 27, February 4 and March 4, 2020 were calculated. Huanggang, Jingzhou and Xiaogan cities were used because the migration scale index of these cities in Hubei province during the same period ranked secondary to Wuhan. There may be potential risk of COVID-19 spread in Huanggang, Jingzhou and Xiaogan cities through migration. Sankey maps of migration from Wuhan, Huanggang, Jingzhou and Xiaogan cities to top 20 cities for the five sub-periods were obtained using ECHARTS 4.7.0.

For each city in China, migration scale index (M_index_) was calculated as follows:

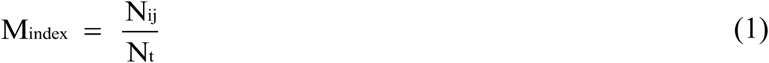

Where M_index_ denote migration scale index, and N_ij_ denotes the number of population migrated from departure city i to destination city j, and N_t_ denotes the total number of migration from departure city i.

To assess the effects of socioeconomic levels on COVID-19 treatment across China, we examined the correlation between socioeconomic levels and confirmed cases, cured cases and the rate of severe cases at the city and province levels (with an α value of 0.05). Hubei province was excluded. Economic levels related to health care were described including population, total health expenditure (billion Yuan), GDP (billion Yuan), total health expenditure in GDP (%), the number of hospitals, the number of top hospitals, the number of doctors, the number of nurses, the number of beds in general hospitals, and public expenditure (billion Yuan). Spatial patterns of the number of confirmed and cured cases and the rates of severe cases across 31 regions and 292 cities of China as of March 4, 2020 were presented using Arcgis Desktop 10.2. For each province, the rate of severe cases was calculated as follows:

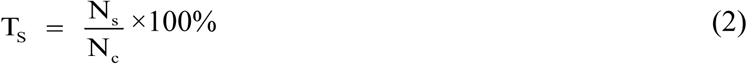

Where T_s_ denotes the rates of severe cases for province i, N_s_ and N_c_ denote the number of severe and confirmed cases in province i, respectively. Here, the number of severe cases consisted of severe and critical cases. Hubei province were excluded from the calculation of the rate of severe cases. The average rate of severe cases in the provinces, municipalities and autonomous regions outside Hubei Province from January 21 to March 7, 2020 was calculated. Temporal changes in the rates of severe cases were obtained using EXCEL 2013.

## 3. Results

### 3.1 Epidemiologic patterns of the COVID-19 outbreak in China

Confirmed COVID-19 cases indicates an exponential growth by human to human transmission since late December, 2019, and a slight decline from January 9, 2020. This decline was not an inflection point but due to the time lag between infection and diagnosis. An average incubation period was 1 to 14 days ^19^, and the COVID-19 suspected cases were diagnosed by viral nucleic acid testing. The first epidemic peak occurred on January 8, 2020, and the cases focused in Wuhan. Starting on January 20, 2020, the case number indicates an explosive increase nationwide, and the second epidemic peak occurred on January 27. On 31 January, a total of 875 confirmed cases, 0.07/100,000 person all over China outside Hubei Province was shown. The case number continued to boom, and the third epidemic peak occurred on February 4 (3156 laboratory-confirmed cases, 5.33/100,000 person in Hubei Province). Afterwards, the cases gradually reduced. Notably, on February 12 and 13, the case peak was not real but due to the number of clinically diagnosed cases was included in Hubei Province. As of February 24, the cases were reported across 319 cities of 34 provinces, municipalities and autonomous regions in China. After Wuhan lockdown, the number of cases reached peak in 12 days, and the COVID-19 pandemic was basically contained in China in four weeks (Figure 1).

Temporal and spatial patterns of suspected and confirmed cases at a province level show that outside Hubei Province, top five provinces in the number of confirmed cases were presented in a decreasing order: Guangdong, Henan, Zhejiang, Hunan and Anhui Province. Top five provinces in the growth rates of confirmed cases were indicated in a decreasing order: Zhejiang, Guangdong, Henan, Hunan and Jiangxi Province (Figure 1). Temporal and spatial patterns of confirmed cases at a city level show that COVID-19 continued to spread from Wuhan to the whole China (Figure 2). Regarding connection characteristics, COVID-19 cases had apparent geographical agglomeration. Wuhan was the single center of agglomeration, and the cases gradually reduced with the increase of geographical distance within a certain range. The cases in Wuhan had an extremely strong connection with those in other cities of Hubei, and had the largest radiation range in Hubei. Additionally, the cases in Wuhan had a strong radiation range in other provinces, and indicate a strong connection with the neighborhood cities and the major cities in other provinces. Regarding the grades of the case number, the differences in the grade gradients between cities were obvious.

### 3.2 Effect of social management measures and migration on the COVID-19 spread in China

A total number of 4.3 million people left from Wuhan between January 11 and 23, 2020^11^. On January 23, Wuhan was closed, by that time, approximately 5 million people had already left for hometowns for the Chinese Lunar New Year, for holidays, and for keeping away from COVID-19^20^. From January 1 to 10, 2020, a total number of 0.7 million people was assessed to leave from Wuhan. Migration from Wuhan indicates an abnormal increase since January 1, 2020 in comparison with that during the same period in 2019. Migration from other cities in Hubei does not indicate an abnormal increase (Figure 3). The migrations from Wuhan in five sub-periods (Figure 3 and 4) were associated with social management measures (Figure 1). Specifically, the first sub-period was for COVID-19 onset between January 1 and 8, 2020. WHC declared COVID-19 outbreak and Huanan Seafood Market was closed on December 31, 2019 and January 1, 2020, respectively. On January 8, 2020, COVID-19 was identified as the pathogen. Due to fear of COVID-19, migration from Wuhan indicates an abnormal increase in comparison with that during the same period in 2019. The second sub-period was for the first COVID-19 epidemic peak between January 8 and 15, 2020. During the period, China CDC started public emergency response level to the highest level. Migration from Wuhan indicates a mild fluctuation, however still an abnormal increase in comparison with that during the same period in 2019. Migration from Wuhan reached the first peak on January 8, 2020 due to increasing fear of COVID-19. The third sub-period between January 16 and 20, 2020 was for the COVID-19 turning phase from Wuhan to the whole China. During the period, COVID-19 was identified as a statutory B infectious disease in China. On January 20, human to human transmission was confirmed, and population started to flee from Wuhan. The fourth sub-period was for the COVID-19 outbreak across China between January 21 and 24, 2020. Migration indicates a sharp increase in comparison with that during the same period in 2019, and peaked on January 23, which was the date Wuhan was closed. The fifth sub-period was between January 24 and 28, 2020. During the period, other cities in Hubei were closed. Migration from Wuhan and other cities in Hubei gradually fell to zero.

**Figure 3.**
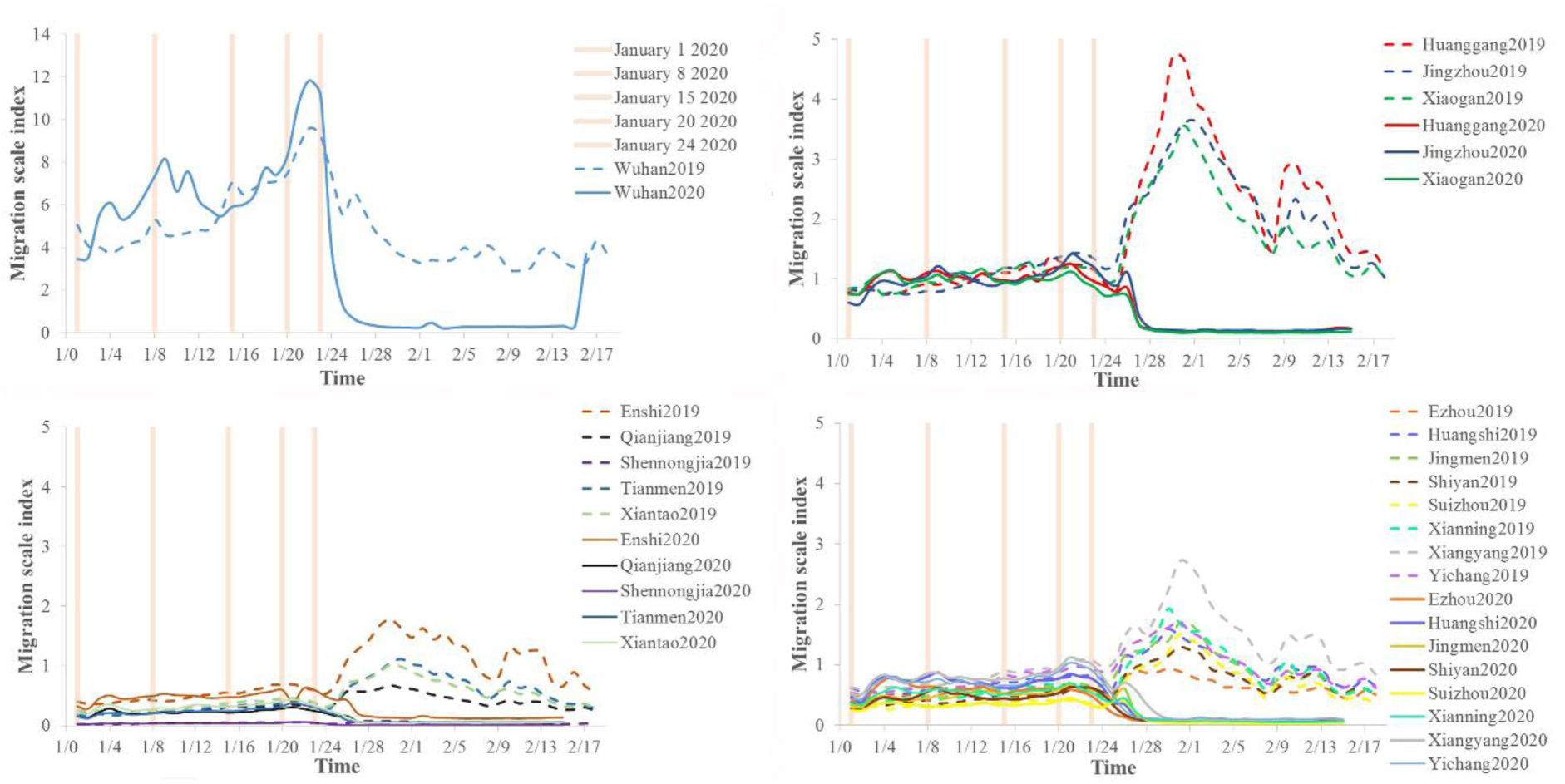
Daily migration scale index of the cities in Hubei Province from January 1 to February 17 in 2019 and 2020. The solid lines denote the migration scale index in 2020, and the dotted lines denote that index in 2019.

The average migration scale index of Wuhan in the five sub-periods were 5.4, 6.5, 7.2, 11.2, and 1.3, respectively (Figure 3). As shown in Figure 4, top 20 hot destination cities that migrated from Wuhan for these sub-periods were Changsha, Chongqing, Beijing, Xinyang, Shanghai, Zhengzhou, Guangzhou, Chengdu, Shenzhen, Jiujiang, Yueyang, Nanyang, Changde, Zhumadian, Nanchang, Nanjing, Hefei, Dongguan, Fuyang, and Zhoukou. These cities were also the top destination cities that migrated from other cities in Hubei for the same period. Migration from Wuhan had a strong radiation range in other provinces, and indicates a strong connection with the neighborhood cities and the major cities in other provinces. Spatial patterns of migration in Wuhan and other cities in Hubei were consistent with those of confirmed cases outside Hubei. A large number of COVID-19 carriers gradually migrated from Wuhan during the five sub-periods, and many people in other cities in Hubei, other provinces, municipalities or autonomous regions of China were infected by close contact with these migration. The cases that occurred in December 2019 were likely to be a small-scale exposure transmission mode in Wuhan, and since January 2020, the cases were likely to be a spread transmission mode in China.

**Figure 4.**
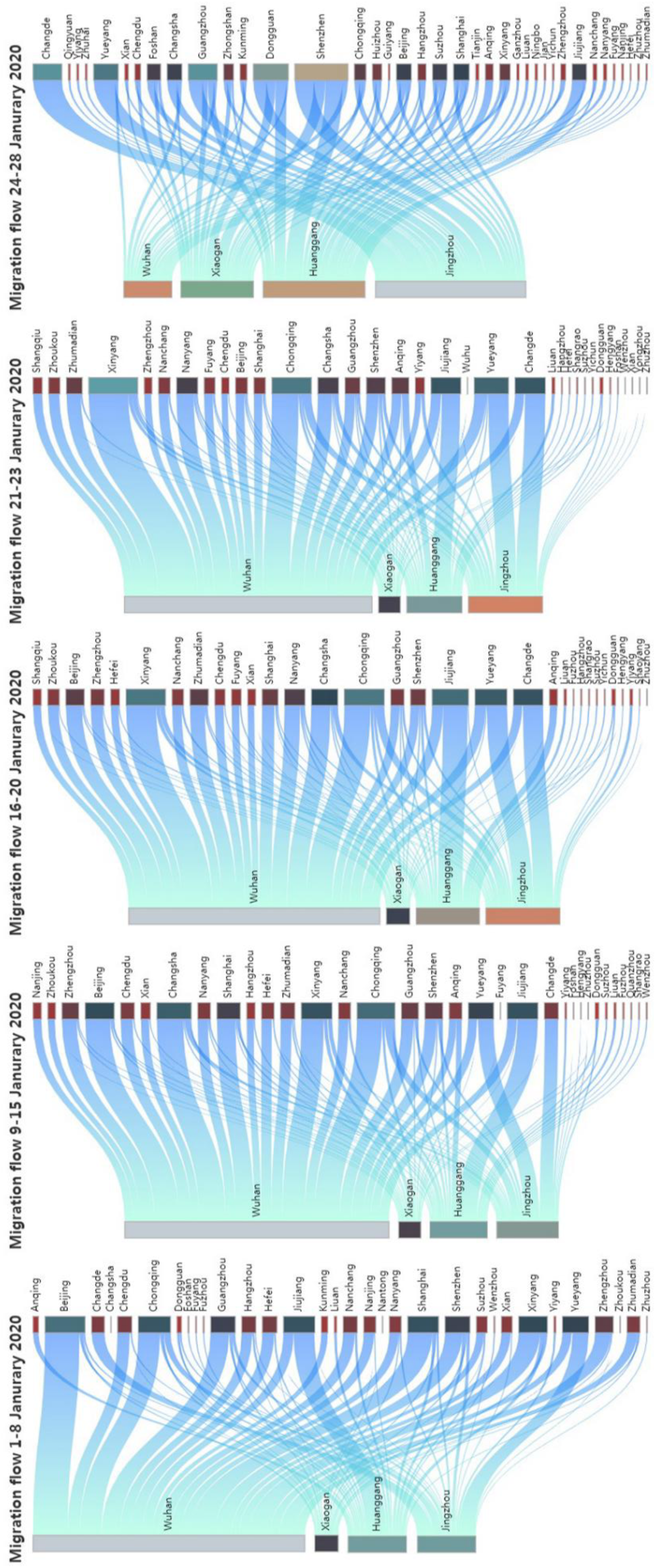
Migration flow on top 20 hot destination cities migrated from Wuhan, Huanggang, Xiaogan and Jingzhou cities of Hubei Province in five sub-periods: January 1 to 8, January 9 to 15, January 16 to 20, January 21 to 23 and January 24 to 28 in 2020.

Temporal patterns of migration in Wuhan, Huanggang, Jingzhou and Xiaogan cities in Hubei indicate significantly positive correlation with those of the case number outside Hubei of China (Table 1). Migration scale indices between January 1 and 8, 9 and 15, 16 and 20, 21 and 23 from Wuhan were significantly correlated with confirmed cases outside Hubei on January 27, February 4 and March 4 (p<0.01) (Table 1). Among the correlations, migration scale indices between January 1 and 8, 9 and 15 from Wuhan were best correlated with confirmed cases outside Hubei on January 27, and the correlation coefficients r were 0.75 and 0.76, respectively (p<0.01). Migration scale indices between January 24 and 28 from Huanggang, Jingzhou and Xiaogan cities indicate significant correlations with confirmed cases outside Hubei. The correlations between confirmed cases and migration were found to be a time lag of approximately 20–30 days or even 50–60 days, while the incubation period of COVID-19 was estimated to be 1–14 days in general^19^. Thus, the COVID-19 outbreak outside Hubei of China was delayed due to the effects of a series of social management measures. The analysis above suggests that since January 1, 2020, 23 days before Wuhan lockdown, the COVID-19 spread that affected by social management measures and migration started across China. The low level of peak incidence per capita, the early timing of the peak, and the effective control of case number, suggest that management measures were associated with a delay in COVID-19 outbreak and a notable decline in case number in China.

**Table 1.** Correlation between migration scale indices from Wuhan, Huanggang, Jingzhou and Xiaogan cities and confirmed cases outside Hubei Province of China

|  | Cases 1.27.2020 | Cases 2.4.2020 | Cases 3.4.2020 |
| --- | --- | --- | --- |
| Migration between January 1 and 8 from Wuhan | 0.75** | 0.70** | 0.71** |
| Migration between January 9 and 15 from Wuhan | 0.76** | 0.70** | 0.70** |
| Migration between January 16 and 20 from Wuhan | 0.67** | 0.62** | 0.63** |
| Migration between January 21 and 23 from Wuhan | 0.51** | 0.52** | 0.56** |
| Migration between January 24 and 28 from Xiaogan | 0.43** | 0.43** | 0.48** |
| Migration between January 24 and 28 from Jingzhou | 0.15* | 0.35* | 0.38* |
| Migration between January 24 and 28 from Huanggang | 0.16* | 0.29* | 0.30* |

\*\*0.01 significance level
\*0.05 significance level

### 3.3 Effect of socioeconomic levels on the COVID-19 treatment in China

Outside Hubei Province, confirmed and cured patients were significantly positively correlated with population, GDP, public expenditure, the number of hospitals, hospital beds and doctors at a city level (p<0.01) (Table 2). At a province level, confirmed and cured cases were well positively correlated with total health expenditure and the share in GDP, GDP, public expenditure, the number of hospitals, top hospitals, beds in general hospitals, nurses and doctors (p<0.01) (Table 3). Spatial patterns of confirmed and cured patients indicate good spatial consistency with those of population, GDP, public expenditure, the number of hospitals, hospital beds and doctors at a city level (Figure 7). Similarly, spatial patterns of confirmed and cured patients indicate good spatial consistency with those of total health expenditure and the share in GDP, GDP, public expenditure, the number of hospitals, top hospitals, beds in general hospitals, nurses and doctors at a province level (Figure 7 and 8).

**Table 2.** Correlation between socioeconomic levels and treatment capacities of COVID-19 at a city level in China.

| City | Population | GDP (billion Yuan) | Public expenditure (billion Yuan) | No. of hospitals | No. of hospital beds | No. of doctors |
| --- | --- | --- | --- | --- | --- | --- |
| Cured | 0.66** | 0.68** | 0.67** | 0.71** | 0.67** | 0.68** |
| Confirmed | 0.67** | 0.71** | 0.70** | 0.73** | 0.70** | 0.72** |

\*\*0.01 significance level
\*0.05 significance level

**Table 3.** Correlation between socioeconomic levels and treatment capacities of COVID-19 at a province level in China.

| Province | Total health expenditure<br>(billion Yuan) | Total health expenditure<br>in GDP (%) | No. of hospitals | No. of top hospitals | No. of doctors | No. of nurses | No. of beds in general hospitals | GDP<br>(billion Yuan) | Public expenditure<br>(billion Yuan) |
| --- | --- | --- | --- | --- | --- | --- | --- | --- | --- |
| Cured | 0.66** | 0.50** | 0.47* | 0.47** | 0.67** | 0.68** | 0.64** | 0.65** | 0.58** |
| Confirmed | 0.72** | 0.52** | 0.52** | 0.52** | 0.72** | 0.73** | 0.68** | 0.70** | 0.64** |
| Rate of severe cases | -0.37* | 0.16 | -0.16 | -0.31 | -0.35* | -0.38* | -0.26 | -0.33* | -0.31 |

\*\*0.01 significance level
\*0.05 significance level

The rate of severe cases was negatively correlated with total health expenditure, GDP and the number of doctors and nurses (p<0.05) (Table 3). Spatial pattern of the severe rates indicates good spatial consistency with that of total health expenditure, GDP and the number of doctors and nurses at a province level. Top average rates of severe cases were presented in a decreasing order: Tianjin city (26.9%), Xinjiang (22.6%), Anhui Province (19.2%), Hunan Province (14.5%), Heilongjiang Province (14.2%), Liaoning Province (13.7%) and Qinghai Province (13.3%). The rates of severe cases in these regions were close to or exceeded 15%. Among these regions, Tianjin city, Xinjiang and Heilongjiang Province had relatively large rates of death on confirmed cases, with the values of 3/136 person, 3/23 person and 13/500 person, respectively. Xinjiang had the highest rate of death on confirmed cases and the second high rate of severe cases in China (Figure 5). The regions with high rates of severe cases were also the regions that invested the least fund in health care in China. Qinghai Province, Tianjin City, Xinjiang, Gansu Province and Hainan Province that were equipped with the least number of doctors and nurses at a province level in China, had relatively high rates of severe cases. Particularly in Qinghai province, the number of doctors and nurses were the least in China, with the values of 13725 and 17577, respectively. Qinghai Province also had the least GDP among all regions in China.

**Figure 5.**
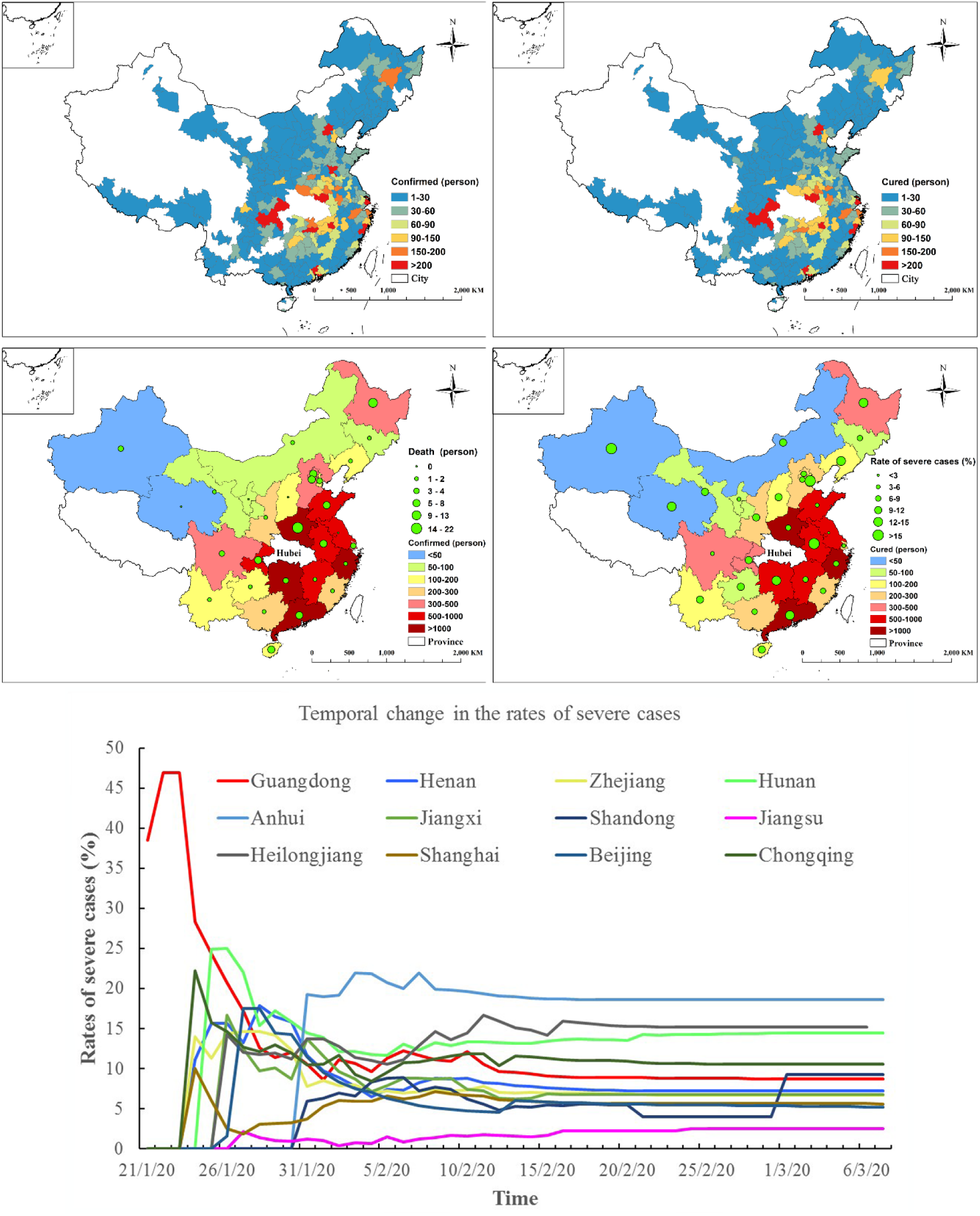
Spatial patterns of confirmed and cured cases at city and province levels, the rates of severe cases and the number of death at a province level outside Hubei of China. Temporal change in the rates of severe cases in the regions with more than 500 confirmed cases.

Considering the number of confirmed, cured and death cases and the rates of severe cases, the regions with the most confirmed cases (more than one thousand) were presented in a decreasing order: Guangdong Province, Henan province, Zhejiang Province and Hunan Province. Zhejiang and Guangdong showed the most excellent performance in COVID-19 treatment capacity. The highest rate of cured cases on the confirmed (96.3%), the lowest average rate of severe cases (7.7%), and the fewest death, only one person in Zhejiang were indicated. Guangdong achieved the largest decline in severe case rate from 31.8% to 9.7% in China. The regions with the confirmed case number between 500 and 1000 were presented in a decreasing order: Anhui Province, Jiangxi Province, Shandong Province, Jiangsu Province, and Heilongjiang Province. Notably, Jiangsu had the lowest average rate of severe cases (1.7%) and zero death among all regions in China. Guangdong, Jiangsu and Zhejiang Provinces, which ranked the top three GDP in China, showed the most excellent treatment capacity of COVID-19 (Figure 5). These three provinces were equipped with relatively more abundant health care resources and investment, and provided the most health care assistance to Hubei. The difference in the number of health care workers that assisted Hubei roughly illustrates the difference in health care levels among regions in China (Figure 6).

**Figure 6.**
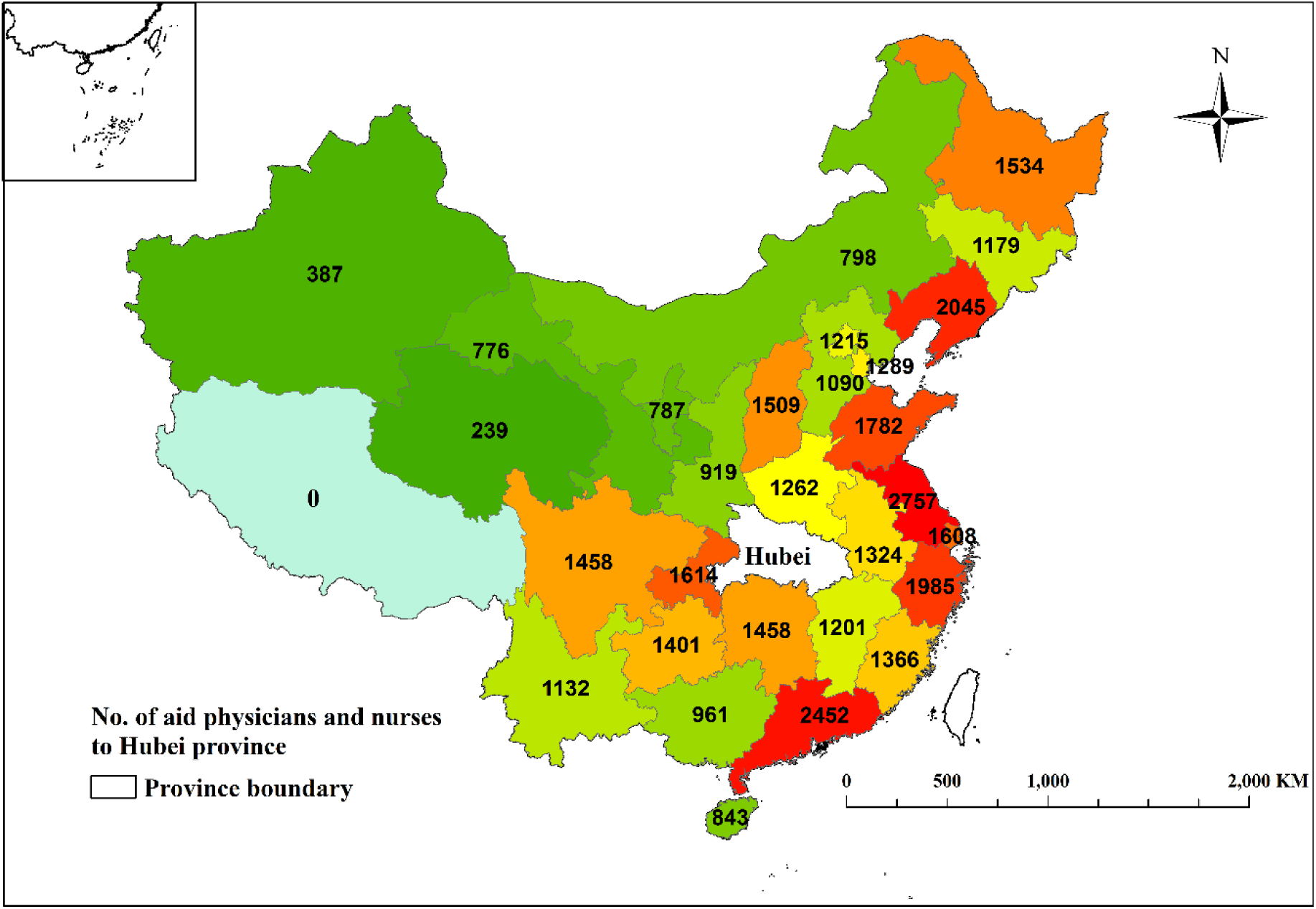
Number of aid physicians and nurses to Hubei Province from other provinces, municipalities, and autonomous regions in China

**Figure 7.**
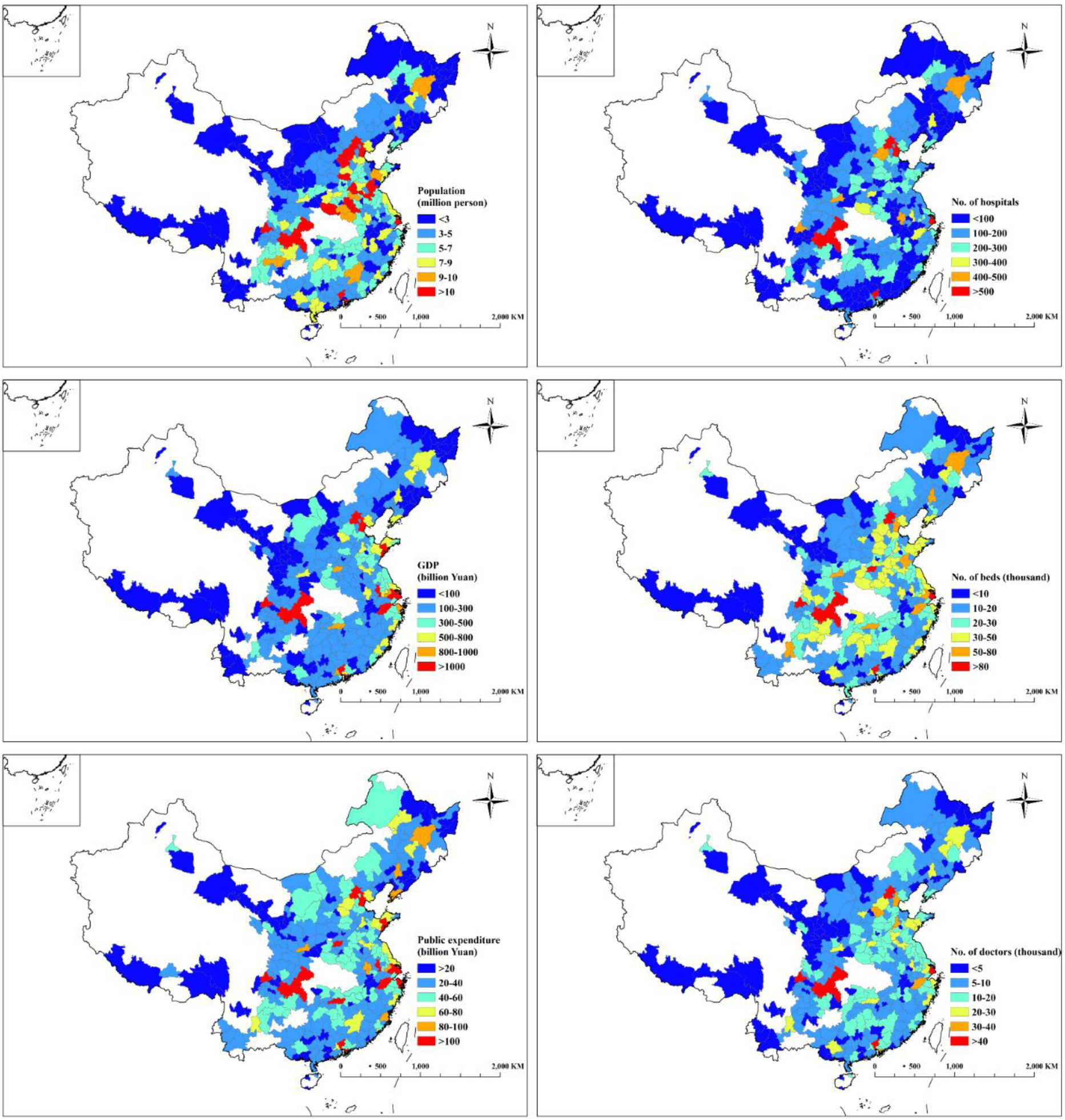
Spatial patterns of the indicators representing macroeconomic levels at a city level in China. The indicators comprised population, GDP (billion Yuan), No. of hospitals, No. of doctors, No. of hospital beds, and public expenditure (billion Yuan).

**Figure 8.**
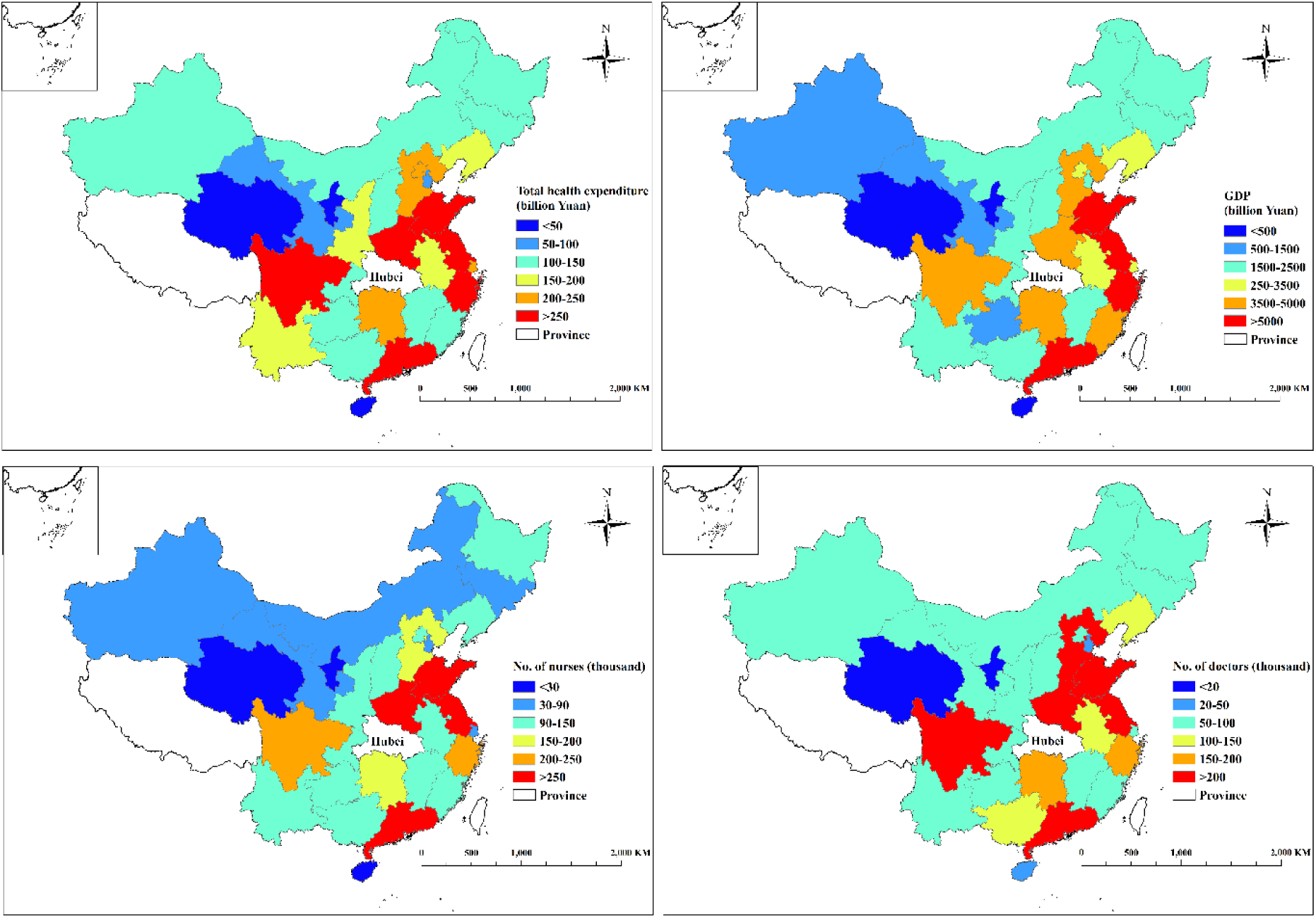
Spatial patterns of the indicators representing macroeconomic levels at a province level in China. The indicators comprised total health expenditure (billion Yuan), GDP (billion Yuan), No. of doctors and No. of nurses.

**Figure 9.**
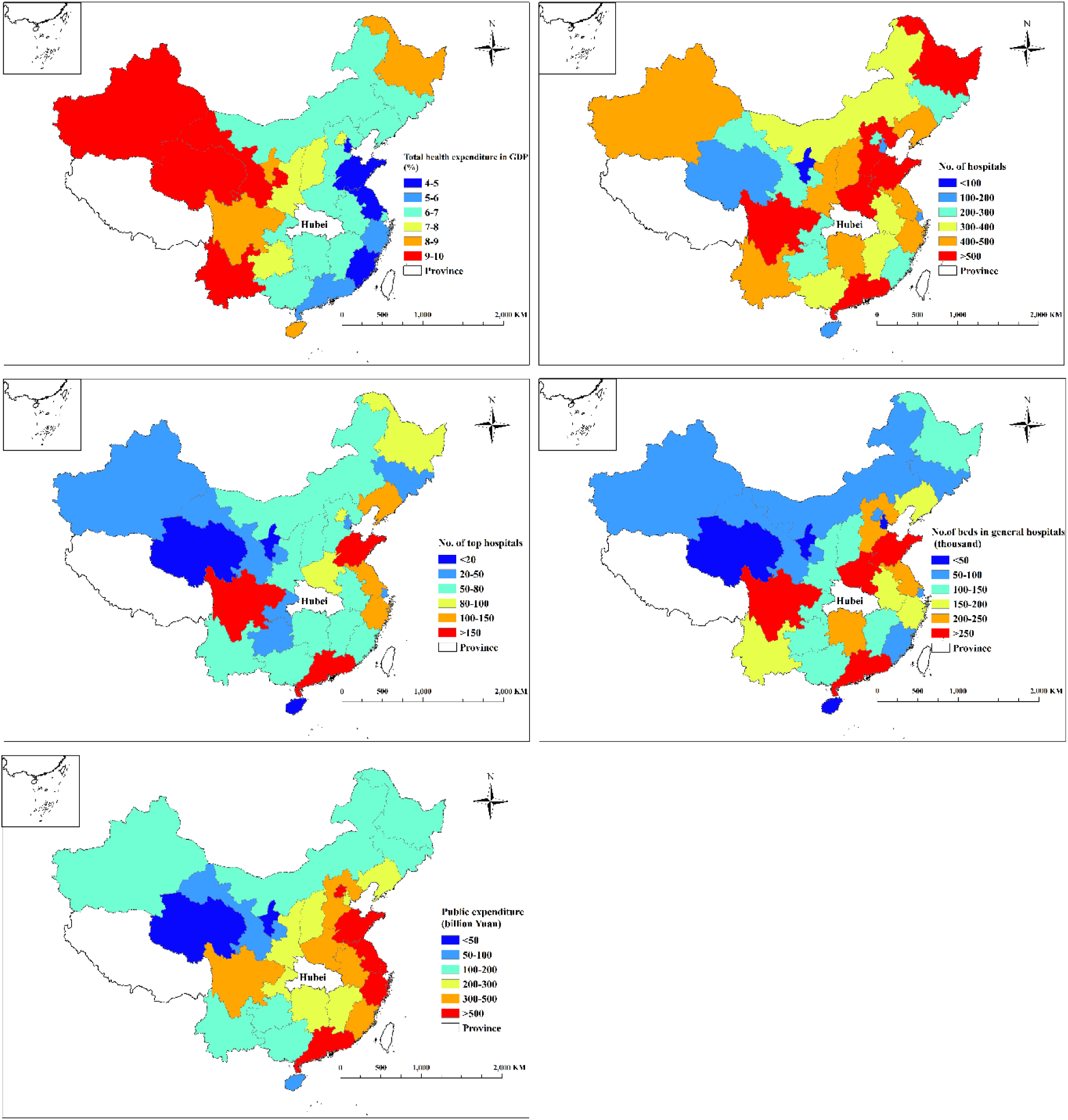
Spatial patterns of the indicators representing macroeconomic levels at a province level in China. The indicators comprised population, total health expenditure in GDP (%), No. of hospitals, No. of top hospitals, No. of beds in general hospitals, and public expenditure (billion Yuan).

## 4. Discussions

### 4.1 Effect of the socioeconomic levels on COVID-19 spread in China

Our results indicate that migration and management measures had a powerful impact on the COVID-19 spread in China. Affected by a series of measures against COVID-19, e.g., the announcement of a new virus outbreak, etiology identification, upgrade of public health emergency level and confirmation of human to human transmission, an abnormal migration from Wuhan occurred between January 1 and 23, 2020 due to fear of COVID-19. Temporal and spatial patterns of migration from Wuhan were consistent with those of COVID-19 cases outside Hubei. Starting on January 23, 2020, affected by management measures, e.g., Wuhan lockdown, movement restriction and home stay throughout China, population mobility and gatherings were reduced, thus containing COVID-19 spread in China. Additionally, Chinese government launched a series of more intensive measures, e.g., wearing masks in public, nationwide joint prevention and control to screening at a community level, four early strategies, and information disclosure. These measures at various levels in China were superimposed, and effectively controlled COVID-19 spread within four weeks after Wuhan lockdown. The findings of Tian, Liu ^11^ based on modelling were similar to our results based on observational data, and they found that the national emergency response appears to have delayed the growth and limited the size of COVID-19 epidemic in China. With the rapid COVID-19 spread, the United States, Italy and Spain is becoming the epicenters. At present, the countries with serious COVID-19 epidemic also adopted intensive measures, e.g., lockdown, wearing masks in public, social distancing, detection and information disclosure, to control and reverse COVID-19 ^21^.

### 4.2 Effect of the socioeconomic levels on COVID-19 treatment in China

Our results suggest that social management measures on monitoring, communication, response, research, epidemiologic survey and clinical practice adopted in China improved COVID-19 treatment. For example, four early strategies, early protection (social distancing), early detection, early diagnosis and early isolation, were implemented across China. As the COVID-19 outbreak threatened to overload the healthcare system and disrupt the global socioeconomic system. The aim of four early strategies is not only to reduce the case number but also to spread them over time, avoiding congestion in health care system and intensive care units. As is often the case during this type of pandemic, health care workers themselves are particularly exposed to infection. Between mid-January and mid-February in China 1716 health care workers were infected with COVID-19 (accounting for 3.8% of all patients)^17^. Four early strategies, Fangcang shelter hospitals, medical assistance nationwide, and continuously updated diagnosis and health care plan for COVID-19 were together carried out. These strategies greatly improved COVID-19 treatment capacities in China, particularly in Hubei Province. Specifically, confirmed, suspected patients and close contacts of confirmed patients were identified, and these people were classified and managed in a centralized way. A large number of mild cases were thus treated timely, and the possibility of turning into severe cases was reduced^22^. These strategies laid a good foundation for increasing the cure rate and reducing the mortality rate. The Fangcang shelter hospitals provided basic health care and clinical triage, and mild, moderate and severe patients were differently treated. It is suitable for the management of a large number of patients and the burden on general hospitals was reduced. Time and economic cost of Fangcang shelter hospital construction is low^23^. With the strengthening force of national medical support and treatment experience, health care resources and capacities were greatly improved.

Our results suggest that COVID-19 treatment capacities outside Hubei in China were greatly affected by regional socioeconomic levels. COVID-19 treatment capacities in Guangdong, Jiangsu and Zhejiang Provinces ranked top three in China, which was consistent with the GDP ranking. Strong economic support is the guarantee of health care level. Strong health care level was the foundation of zero death and extremely low rate of severe cases in Jiangsu Province. Despite a large number of COVID-19 confirmed cases (631 cases) in Jiangsu, early detection, early treatment, and rapid recovery were achieved in the context of sufficient healthcare resources, and the rate of severe cases was the lowest in China. Additionally, the total number of medical members from Jiangsu assisting Hubei is 2,757, which was the largest among the regions in China (Figure 6). The Jiangsu medical team took over treatment work in Xiaogan City, Hubei Province, and focused on the intensive care units. The confirmed cases (3518 cases) in Xiaogan was secondary to Wuhan in Hubei Province. In Wuhan, there were five key hospitals that received the severe and critical patients. Jiangsu medical team participated in three of these hospitals as the main force^24^. These aspects manifested powerful treatment capability of Jiangsu Province.

Guangdong was the second largest province to provide medical assistance to Hubei. Nevertheless the number of COVID-19 confirmed cases in Guangdong was the largest, the rate of severe cases in Guangdong decreased the most by 22.1% among the regions in China. This large decrease may be mainly due to continuously improved diagnosis and treatment experience since COVID-19 outbreak. Guangdong established a comprehensive screening system for the key COVID-19 cases. The screening team is composed of medical experts from the Department of Infectious Diseases, Respiratory Medicine, Intensive Medicine, and Imaging. This screen was performed on confirmed patients every day to examine if these patients had the tendency to convert to severe or critical cases and take remedy measures. The number of severe and critical cases was thus gradually declined. Besides, timely and effective publicity measures were conducive to early detection of patients and avoided illness deterioration.

The excellent treatment capacity of COVID-19 in Zhejiang Province was mainly presented from these aspects: early prevention and control, early development of detection reagents, strong detection capability, and centralized treatment strategies. Specifically, Zhejiang started the first level response of public health emergency and took the strictest measures of joint prevention and control at the earliest. On January 10, 2020, Zhejiang Provincial Health Commission carried out hospital-infection arrangements for hypothesized epidemic outbreak in medical institutions across the province. Zhejiang CDC was the first provincial disease control laboratory to successfully isolate COVID-19 coronavirus strains in China, and an automated whole-genome detection and analysis platform was launched to speed up COVID-19 detection. Many institutions in Zhejiang had the detection ability at the initial phase of COVID-19 outbreak, with a daily detection capacity of 12,000 person. At the same time, a third-party testing agency in Zhejiang undertook a total number of 75,000 nucleic acid testing in neighboring provinces by mid-February 2020^25^. With the support of screening at fever clinics and nucleic acid detection, early treatment was achieved. Moreover, four centralization strategies were adopted in treatment in Zhengjiang. Patients and medical resources were centralized, and medical experts and treatment were centralized. Treatment was centralized based on disease severity. The four centralization could save medical resources, improve the treatment efficiency and the rate of cured cases, and reduce the rates of severe cases and mortality.

Regarding COVID-19 detection, the above three provinces showed strong nucleic acid detection capabilities and had the ability to assist the detection in other provinces of China. In China, most of famous enterprises of nucleic acid detection kit development are located in Guangdong, Jiangsu and Zhejiang Provinces, for instance, Jinyu and Huada in Guangdong, Shuoshi and Suzhou Medical Workers in Jiangsu, Dean and Adicon in Zhejiang. To improve detection efficiency, at the beginning of COVID-19 outbreak, a three-party joint test, “CDC + medical institution + the third-party enterprise”, was carried out in these three provinces. The testing results could be produced on the same day, and there was no problem of backlog of suspected patients due to timely diagnosis. Additionally, Tianjin city, Xinjiang and Qianghai Province had higher rates of severe cases and mortality in comparison with other regions in China. Because these regions had a relatively low level of economic investment in health care, and the treatment capacities of COVID-19 were thus relatively poor.

Our key finding is that socioeconomic levels had strong effects on the spread and treatment of COVID-19 in China. Future work on the individual effect of each management measure on the spread and treatment of COVID-19 at a province level could be carried out outside Hubei Province, both quantitatively and qualitatively. The effect of each management measure could be examined separately and the dominant measure could be examined. A comparative analysis using control experiments and model simulation on the effects of these management measures at a province or country scale could be performed. Additionally, the limitation of this study lied in data unavailability. For an improved understanding of socioeconomic effect on COVID-19 treatment in China, case reports of severe and dead patients at a city level, including gender, age, weight, medical history and length of hospital stay could be added. Socioeconomic factors, e.g., the number of beds, doctors and nurses in intensive care units, the number of key medical resources in all top hospitals, e.g., negative pressure ambulances, Extracorporeal Membrane Oxygenation (ECMO), ventilator and other key treatment equipment at a city level could be supplemented. COVID-19 spread in China was effectively curbed, and the spread to other countries was notably reduced. The effectiveness of these Chinese measures in controlling transmission and treatment of COVID-19 in other countries of the world requires intensive examination. The Chinese experience provides important insights into how to design effective management strategies of COVID-19 or other epidemic outside China.

## Data Availability

We collected the COVID-19 cases on a daily basis that reported by the CDCs at all levels as of March 4, 2020, in the cities of 31 provinces, municipals and autonomous regions in Mainland China.These data and management measures were obtained from the websites of local health commissions.Data of migration from Wuhan and the other cities in Hubei Province on a daily basis were obtained from Qianxi Baidu website (https://qianxi.baidu.com/).Macroeconomic data were collected from two levels: city and province level. At the province level, the data from the 31 provinces, municipalities and autonomous regions outside Hubei Province were obtained from China health statistics yearbook in 2019.

https://qianxi.baidu.com/

## Acknowledgements

Pay tribute to the workers who are fighting on the front line of the prevention and control of COVID-19 all over the world. Thank you very much to all the staff who participated in the prevention and control of COVID-19, including treatment, detection and diagnosis, epidemiologic investigation, close contact management, and scientific research, etc.

## Contributors

Qi Wang designed the study concept. Zelong Zheng and Qi Wang collected data and checked data sources. Qi Wang, Xiangfeng Li analyzed data and prepared results. Zelong Zheng wrote the first draft of the manuscript and Qi Wang contributed to subsequent drafts.

